# Sero-prevalence Findings from Metropoles in Pakistan: Implications for Assessing COVID-19 Prevalence and Case-fatality within a Dense, Urban Working Population

**DOI:** 10.1101/2020.08.13.20173914

**Authors:** Wajiha Javed, Jaffer Bin Baqar, Syed Hussain Baqar Abidi, Wajiha Farooq

## Abstract

Population-level serologic testing has demonstrated groundbreaking results in monitoring the prevalence and case-fatality of COVID-19 within a population. In Pakistan, Getz Pharma conducted a sero-prevalence survey on a sample of 24,210 individuals using the IgG/IgM Test Kit (Colloidal gold) with follow-up and sequential testing after every 15-20 days on a sub-sample. This is the first of its kind, large scale census conducted on a dense, urban, working population in Pakistan. The study results reveal that from 24,210 individuals screened, 17.5% tested positive, with 7% IgM positive, 6.0% IgG positive and 4.5% combined IgM and IgG positive. These findings have been extrapolated to the rest of the urban, adult, working population of Pakistan, and as of 6th July, 2020, 4.11 million people in Pakistan have been infected with COVID-19, which is 17.7 times higher than the current number of 231,818 symptom-based PCR cases reported by the government which exclude asymptomatic cases.

## Introduction

Serologic tests are based on the qualitative detection of IgM and IgG generated by the body in response to a SARS-CoV-2 infection.^1^ IgM is the first antibody type generated by the body in response to a COVID-19 infection, followed by IgG which replaces IgM as the predominant antibody in the blood.^1^ Given that Pakistan is facing an issue of limited testing capacity focused exclusively on symptom-based PCR tests, there is a need to conduct sero-prevalence studies to determine the true extent of the disease via serologic tests. According to a paper published in a peer-reviewed journal in JAMA, even the "gold standard" PCR kits can have an accuracy ranging from 32% (pharyngeal swab) to 63% (nasal swab) depending on the technique of sample collection, the site and the phase of the disease.^2^

Although literature indicates that asymptomatic infections within a population may be many folds higher than the number of PCR reported cases,^3,4^ large-scale, sero-prevalence studies within low-risk populations have not yet been conducted in Pakistan. Hence, in order to expand the database of COVID-19 infections in the country, Getz Pharma conducted a random testing (census) of 24,210 individuals from various workplaces in metropoles of Pakistan, using USFDA EUA and CE approved serologic test kits. The kits are currently being used in 38 countries globally, as they have a sensitivity of 95.3% and specificity of 98.7% for IgG, and a sensitivity of 86.48% and specificity of 95.18% for IgM,^5^ with a false positive rate ranging from 2% to a maximum of 7%,^3^ due to nonspecific immunity and nonspecific protection to COVID-19. Hence, serologic testing kits can be used as cost-effective measures for conducting mass screening amongst asymptomatic individuals, who would otherwise not present at a government-approved PCR testing facility. Consequently, the official number of confirmed cases severely under-represent the full extent of COVID-19, as they fail to capture the proportion of the population who are asymptomatic carriers and are actively spreading the infection to more vulnerable members of society.

## Methods

Getz Pharma conducted a 100% census sero-prevalence study on a sample of 24,210 individuals using the IgG/IgM Test Kit (Colloidal gold) with follow-up and sequential testing after every 15-20 days. The sample size included an adult, working population aged 18-65 years, recruited from dense, urban workplaces including factories, corporates, restaurants, media houses, schools, banks, healthcare providers in hospitals, and families of positive cases in various metropoles of Pakistan (Karachi, Lahore, Multan, Peshawar and Quetta). This was a large-scale, cross-sectional study conducted amongst the general, low-risk population. While extrapolating the study findings to the general population of 220 million people in Pakistan, 53% of the population which is under 18 years of age and 4.5% above 65 years needs to be stratified and excluded, thus restricting the universe to 93 million. Similarly, the urban population setting of this census excludes 64 percent of the rural population of Pakistan, thus reducing the universe to 33 million individuals from the urban, adult working population. It must be noted that the infectivity quotient (R0) of COVID-19 is very different between densely urban and less crowded, rural populations ranging from 1.4 to 3.9.^6,7^

## Results

The study results revealed a total of 17.5% COVID-19 positive cases from a sample size of 24,210 individuals. Most of these were ongoing infections at 11.5%, while 6% had recovered. From the sample of 24,210 individuals recruited in the study, a total of 8,937 registered employees were screened from factories and corporate offices. Out of these, 15.2% tested positive. Specifically, 7.2% tested IgM positive, while 4.8% tested IgG positive and 3.2% were combined IgM and IgG positive. This prevalence can be extrapolated to the one million registered working population of Karachi, meaning at least 152,000 infected cases in Karachi alone, with 104,000 being currently exposed, unaware and spreading infection to people around them. These findings can be applied to the remaining urban workforce of Pakistan with similar demographics, between the ages of 18-65 years. By taking a base population of 61.7 million registered workers^8^ within this age range, assuming that 36% live in urban areas with similar workplace dynamics (22.21 million), it can be extrapolated that 4,110,381 (4.11 million) from the working population are currently infected with COVID-19 as of 6^th^ July, 2020. From a total of 896 individuals screened from media houses and print media, 8.6% tested positive, with 4.7% IgM positive, 2.9% IgG positive and 1% with both. Taking a base population of 50,000 media individuals in Pakistan including mainstream and print media, we can extrapolate that 4,297 individuals from the media industry of Pakistan are currently exposed to COVID-19. Amongst 3,120 healthcare workers including doctors and paramedics from different metropoles in Pakistan, 17% tested positive, with 4.1% currently infected and 4.6% IgG positive which means that they had been infected in the past and have now recovered. Taking a base population of 313,457 healthcare workers^9^ across Pakistan as per WHO EMRO, we can extrapolate that 53,248 healthcare workers are currently exposed to COVID-19. Out of a total of 7,857 individuals screened who were household contacts of a positive case, it was found that 15.9% individuals tested positive, with 4.2% IgM positive, 4.8% IgG positive and 6.8% with both. With 231,818 PCR cases being reported as of 6^th^ July, 2020^10^ and taking an average Pakistani household size of 6.7^11^, the base population on family members of positive cases is 1,553,181. Given a 15.9% secondary household attack rate, we can extrapolate that 246,508 household members of positive cases are currently exposed to COVID-19. Out of a total of 3,400 symptomatic individuals in the study who requested for symptom-based testing at their households, 30% tested positive, with 16% IgM positive and 14% IgG positive. As of 6^th^ July, 2020, there have been 1,420,623^10^ symptom-based COVID-19 tests across Pakistan. If the study was to be extrapolated to all tests done who presented with symptoms, given the false negativity of PCR especially when viral load is less, with a 30% prevalence of test positivity amongst individuals who have symptoms, at least 426,187 people currently have COVID-19 in Pakistan. Overall, from a sample of 24,210 individuals screened, 17.5% tested positive, with 7% IgM positive, 6.0% IgG positive and 4.5% combined IgM and IgG positive. Given the above extrapolations while keeping the study limitations in mind, we can extrapolate that as of 6^th^ July, 2020, currently 4.11 million individuals in Pakistan have been infected with COVID-19 as opposed to 231,818 cases that the official figures of Pakistan are quoting.^10^ (Table 1)

**Table 1:** Seroprevalence study results and extrapolations.

|  | No. of Screened Individuals | Total Positives |  | IgM |  | IgG |  | Both IgM/IgG |  | Base population used for extrapolation | Extrapolated number of COVID-19 cases in Pakistan |
| --- | --- | --- | --- | --- | --- | --- | --- | --- | --- | --- | --- |
|  | n | n | n% | n | n% | n | n% | n | n% |  |  |
| Corporates /factories | 8,937 | 1,360 | 15.2% | 646 | 7.2% | 425 | 4.8% | 289 | 3.2% | 22,212,000 | 3,380,141 |
| Mainstream Media and print media | 896 | 77 | 8.6% | 42 | 4.7% | 26 | 2.9% | 9 | 1.0% | 50,000 | 4,297 |
| Health care worker | 3,120 | 530 | 17.0% | 128 | 4.1% | 143 | 4.6% | 259 | 8.3% | 313,457 | 53,248 |
| Secondary Household attack rate among positives | 7,857 | 1,247 | 15.9% | 331 | 4.2% | 381 | 4.8% | 535 | 6.8% | 1,553,181 | 246,508 |
| Tests among symptomatic population | 3400 | 1,020 | 30.0% | 544 | 16.0% | 476 | 14.0% | 0 | 0.0% | 1,420,623 | 426,187 |
| Overall | 24,210 | 4234 | 17.5% | 1691 | 7.0% | 1451 | 6.0% | 1092 | 4.5% |  | 4,110,381 |

IgM: Immunoglobulin M
IgG: Immunoglobulin G

## Discussion

Getz Pharma’s sero-prevalence study revealed that 90% of the population who tested COVID-19 positive (screened via serologic tests) were asymptomatic carriers of the disease, who would otherwise have not presented at a PCR testing facility. This is the first of its kind, large scale census conducted on the general, urban population of Pakistan, which indicates the total number of COVID-19 positive cases is 17.7 times higher than symptom-based PCR reported figures. While the study showed sero-prevalence of 17.5% at workplaces, the newly emergent cases at 6 weeks had an incidence rate of 7%, which reveal groundbreaking findings in the context of the evolving COVID-19 situation in Pakistan. Although the study results are restricted to Pakistan’s dense, urban, adult, working population, it can still provide useful insights for guiding public health practices and formulating policies to minimize the human and economic losses incurred due to COVID-19. Before the findings can be considered conclusive, further studies are required, including a household, multistage, cluster random survey in order to find the true extent of COVID-19.

## Data Availability

We assure that all data captured and presented herein, shall be available as and when needed.

## Acknowledgement

We acknowledge the manufacturer Genrui Biotech Inc for the provision of (2019-nCoV) IgG/IgM Test Kits (Colloidal gold) for this study.

## Disclaimer

As is the case with any epidemiological survey, there is a need for further studies to be conducted before the findings can be considered conclusive.

## References

1. Davis, 2020. How Do the COVID-19 Coronavirus Tests Work? Available from: https://www.medicinenet.com/howdothecovid-19coronavirustestswork/article.htm

2. Wang W, Xu Y, Gao R, Lu R, Han K, Wu G, Tan W. Detection of SARS-CoV-2 in different types of clinical specimens. Jama. 2020 May 12;323(18):1843-4.

3. Bendavid E, Mulaney B, Sood N, Shah S, Ling E, Bromley-Dulfano R, Lai C, Weissberg Z, Saavedra R, Tedrow J, Tversky D. COVID-19 Antibody Seroprevalence in Santa Clara County, California. MedRxiv. 2020 Jan 1.

4. Shakiba M, Nazari SS, Mehrabian F, Rezvani SM, Ghasempour Z, Heidarzadeh A. Seroprevalence of COVID-19 virus infection in Guilan province, Iran. medRxiv. 2020 Jan 1.

5. Xu Wanzhou, Li Wei, He Xiaoyun, Zhang Caiqing, Mei Siqing, Li Congrong. Serum 2019 New coronavirus IgM and IgG antibodies jointly detect the diagnostic value of the new coronavirus infection. J/OL. Chinese Journal of Test Medicine. 2020;43. DOI: 10.3760/cma.j.cn114452-20200223-00109

6. Li Q, Guan X, Wu P, Wang X, Zhou L, Tong Y, et al. (January 2020). “Early Transmission Dynamics in Wuhan, China, of Novel Coronavirus-Infected Pneumonia”. The New England Journal of Medicine. 382 (13): 1199-1207. doi:10.1056/NEJMoa2001316. PMC 7121484. PMID 31995857.

7. Riou J, Althaus CL (January 2020). “Pattern of early human-to-human transmission of Wuhan 2019 novel coronavirus (2019-nCoV), December 2019 to January 2020”. Euro Surveillance. 25 (4). doi:10.2807/1560-7917.ES.2020.25.4.2000058. PMC 7001239. PMID 32019669.

8. Shaikh H. How can we protect contract and informal workers in Pakistan? [Internet]. DAWN.COM. 2019. Available from: https://www.dawn.com/news/1484090#

9. WHO EMRO | Health service delivery [Internet]. Emro.who.int. 2020. Available from: http://www.emro.who.int/pdf/pak/programmes/service-delivery.pdf?ua=1

10. COVID-19 Health Advisory Platform by Ministry of National Health Services Regulations and Coordination [Internet]. Covid.gov.pk. 2020. Available from: http://covid.gov.pk/stats/pakistan

11. Ahmed T, Ali SM. Characteristics of Households and Respondents. Pakistan Demographic and Health Survey. 1992 Jul;1:990-1.

